# Benchmarking foundation models for improving confounding control in target trial emulation

**DOI:** 10.64898/2026.05.09.26352820

**Authors:** Sonja Kleper, Rachel Melamed

## Abstract

Machine learning models for causal inference aim to adjust for confounding factors that are associated with both an exposure and an outcome, creating a spurious biased association. But, these methods are rarely empirically evaluated to assess their success in mitigating such bias. Recent advances in knowledge representation, including both foundation models and knowledge graphs, could enrich these models, but rigorous evaluations are needed in order to assess their potential. Here, we ask whether enriching existing causal inference models with knowledge representations from foundation models can improve confounding control. Rather than using semi-simulated data to address this question, we focus on examples of real confounding: we emulate target randomized active comparator trials that are subject to confounding by indication. Our results can guide researchers aiming to develop or apply methods for discovering causal effects from observational data.

## Introduction

Using observational data to answer causal questions has the potential to accelerate advances in domains such as medicine or public policy. One major application is assessing effects of common medications on disease. While randomized clinical trials remain necessary for quantifying true effects, that approach is not scalable for discovering unexpected drug effects, and findings may not generalize. Therefore, emulation of target clinical trials uses real-world data to test the effect of a treatment on disease. For example, we can ask whether people taking GLP-1s have improved cardiometabolic outcomes (Wang et al. 2023). While a clinical trial testing such a question would randomize treatment assignment, in contrast, in observational data treatment is given as a result of a patient’s medical history. This results in confounding: for example people who are wealthier are more likely to take GLP-1s, and they are more likely to have good cognitive health, creating a spurious protective association.

A number of machine learning models have been developed in order to mitigate this confounding bias, a task called causal inference (Louizos et al. 2017; Shalit et al. 2016; Shi et al. 2019; Ma et al. 2021). Most of these models use tabular data as input. But health records data is comprised of a sequence of medical codes. A number of machine learning models, called foundation models, aim to create computational representations of these codes (Li et al. 2020; Huang et al. 2024; Wornow et al. 2023). There is one example of a causal inference model that incorporates representations of the health record using a foundation model (Rao et al. 2024). But, the impact of using tabular data or knowledge representations has not been well evaluated. Therefore, here we propose to perform a systematic evaluation of the potential for enriching causal inference methods for health records with knowledge graph and foundation models in order to improve confounding control.

Choosing the appropriate evaluation is essential for assessing the ability of a model to overcome confounding. In a high-throughput target trial emulation testing for drug effects on cancer, we showed that even when a wide range of medical variables are included in a linear model, confounding bias persists(Lalagkas and Melamed 2025a). As long as this residual confounding remains, it is difficult to know if any drug effect discovered from observational data is a true effect as opposed to the results of bias. While our results were empirical, typically, causal inference models are evaluated using semi-simulated data. This is because, unlike in most machine learning tasks, true drug effects are not known. Foundation models are typical evaluated using their training tasks, including predicting held out medical events. Neither of these typical types of evaluations directly test whether the models are able to overcome realistic examples of confounding.

Therefore, here we prioritize empirical evaluation of the ability of the models to overcome residual confounding, such as we have uncovered in our previous work. A related effort is known as empirical calibration(Schuemie et al. 2018). In that approach, drug effects are compared to a set of likely null effects. For example, we do not expect that antidepressants change your risk of hangnail. Rather than testing the numerous likely non-causal effects, here we ask the models to perform a more difficult task. We ask: can any of these models overcome likely confounded associations? We curate two examples of confounding, and systematically test a range of causal inference models, foundation models, and knowledge graphs for their contribution to overcoming confounding.

## Methods

### Implementation of target trial emulation

For each of the two target trials, we identify 1) cohorts of people taking either of the active comparator treatments, 2) their relevant health history, and 3) their outcome after drug exposure.

For Step 1 and 2 of the target trial emulation (Table 1), we identify users of medications, and their date of first use, using the All of Us Drugs table. The first date of use of either of the two medications is defined as the index date.

**Table 1:**
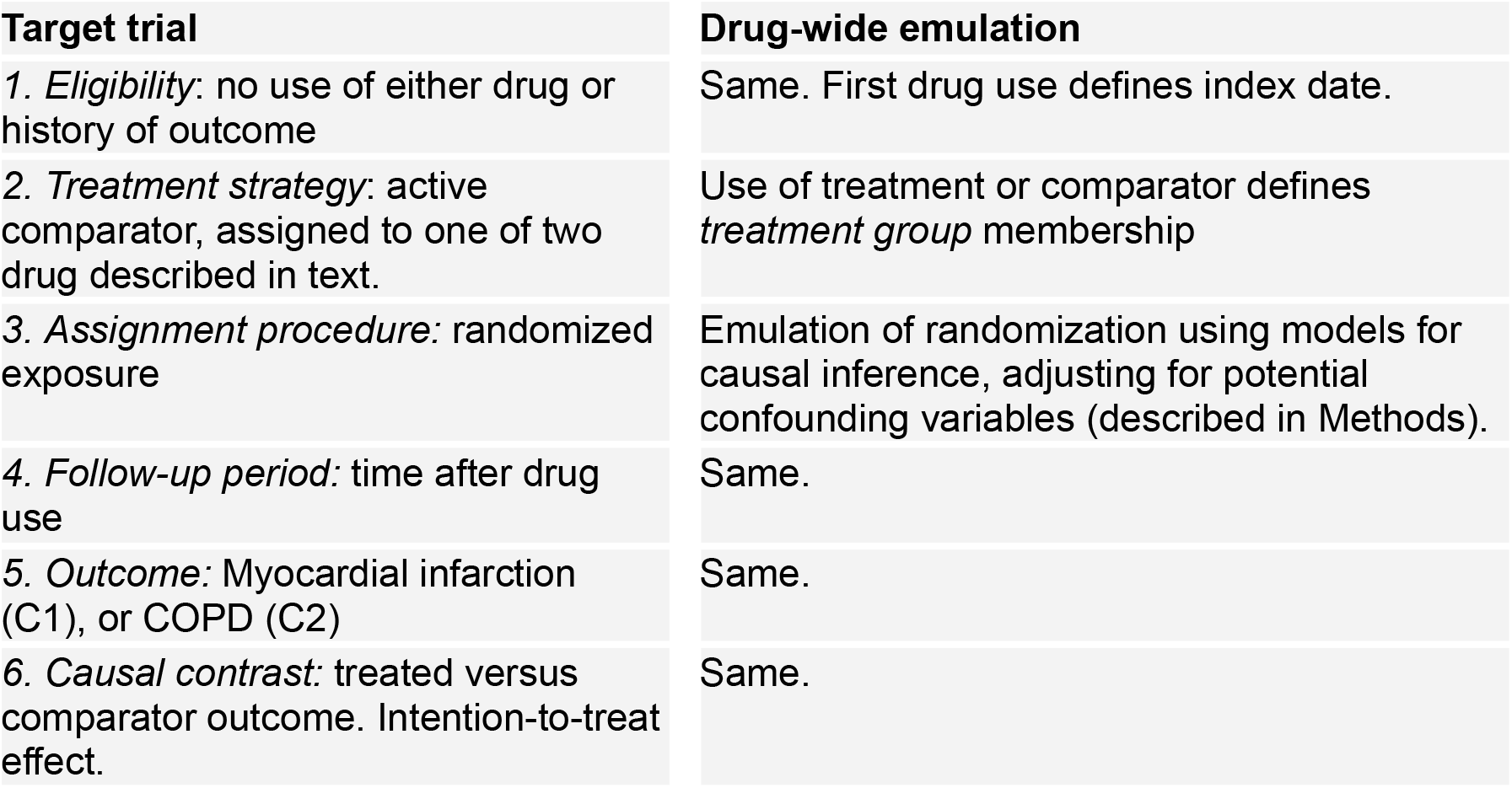
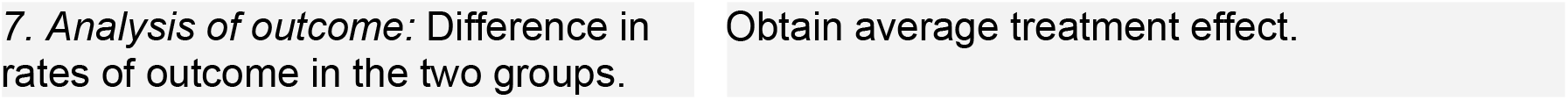
Procedure for emulating the seven components of the target trial.

In order to identify the variables to adjust for in Step 3, for each comparison, we use expert designed cohort studies from literature. For metoprolol versus sumatriptan (C1), we combine information from three studies (Petersen et al. 2024; Dondo et al. 2017; Bruun et al. 2025). For bupropion versus sertraline (C2), we use the designs from two studies(Gaya et al. 2024; Tran et al. 2024). The status of each of these variables was determined from the All of Us observation table, drug table, and condition occurrence tables.

For Step 4 and 5, we follow each individual in the time after the index date, assessing their incidence of respective outcomes using phecodes for first diagnoses in the condition table. Step 6 depends on the causal inference model. For counterfactual outcome models, they are trained to predict two outcomes, estimating their probability of disease if they had taken either of the two drugs 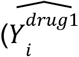 and 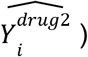. From these quantities we obtain the average difference in counterfactual outcomes, defined as the average treatment effect 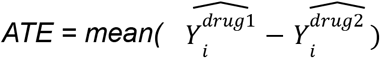. For inverse probability of treatment weighting, models are trained to predict probability of drug, and the probability is transformed into weights using standard methods(Hernán and Robins 2024). Then, the effect of exposure on outcome is assessed in weighted logistic regression.

of the respective depending on the variable.

expert-curated lists We use the health records data available from the All of Us resource to create cohorts for target trial emulation. First, we use the drugs table to identify use of the drugs in our study. Then, we use the condition table and the drugs table to obtain their relevant medical history. and the to antidepressant, and we used the concept relation tables to match antidepressants to their active ingredients, resulting in 18 antidepressants that each had more than 1000 users. For each user we obtained 1) history of factors that might influence severe COVID-19 outcome, collected before positive COVID-19 test, denoted as the set of variables *L*; 2) which antidepressants they used before or at time of COVID-19 diagnosis; 3) presence of severe COVID-19 outcome (defined as severe hospitalization including ventilation, or death), as compared to non-severe COVID-19 outcome (no hospitalization, or hospitalized without ventilation).

Next, we identified cohorts for each of our 153 emulated trials. Each trial compared COVID-19 positive people taking one of two antidepressants. Users taking either of the two drugs were assigned to one of the two arms, and those taking both drugs were removed from the study. We assessed the imbalance between the two treatment groups within a cohort, for each risk factor *L*_*i*_ in *L* as the standardized absolute difference in frequency: 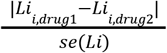. We also calculate the difference in the frequency of severe outcome between the two cohorts.

### Modeling confounding with a range of causal inference models

We implement Tarnet, the S-learners and T-learners using CausalML package(“About CausalML — Causalml Documentation,” n.d.), with both Logistic Regression and multi-layer perceptron base models For logistic regression we used cross validation to select the hyperparameters including l1_ratio (0, 0.25, 0.5, 0.75, or 1) and C. We adapted the code for TarNet from(Koch [2021] 2025). The model was tuned on a learning rate range of: 5e-5, 1e-5, 5e-4, 1e-4, 5e-3, 1e-3, or 1e-2. For the propensity weighting models we similarly use either logistic regression or a multi-layer perceptron to predict propensity. Then, using the propensity weights in a weighted logistic regression we estimate the effect of drug exposure on outcome. In all models, a training set is used for fitting the respective counterfacutal outcome or propensity models; a test set is used for estimating the treatment effect. The confidence intervals are obtained by resampling the held out set with replacement and obtaining the average treatment effects in these bootstrap samples. We also obtain empirical Z-score estimates defined as the ATE divided by two times the bootstrap standard error.

### Representing medical knowledge with foundation models and knowledge graphs

Each medical code representation by passing their names through foundation models and extracting their representation. The models were tuned on biomedical data and selected from the literature. The list is: biobert, biomednlp, biomegatron, bluebert, clinical kb, sapbert, and biogpt. For encoder models (BeRT), the final token is used as the representation. This is because bert-like models have bidirectional attention and are typically pretrained with a prepended [CLS] token which signals the beginning of a sentence, as well as a summary of it. For causal models (GPT), left-to-right attention is used so there is no summary token. Instead, the final token is used as it has seen the full context from the rest of the tokens.

Each medical code in the health record is mapped to connections in two knowledge graphs. The first knowledge graph is derived from the Observational Medical Outcomes Partnership concept relation table, a table relating medical codes to each other. This resources is itself compiled from a number of other manually created databases, including from RxNorm, Snomed, and UMLS. The second knowledge graph is called ClinGraph, which similarly combines multiple knowledge bases.

### Evaluation of knowledge graph and foundation models

We evaluate the consistency between knowledge graphs and foundation language models by mapping each language model embedding to the nodes of the knowledge graph as described above. Then, we ask whether neighboring nodes have similar embeddings, as measured in the cosine similarity. We take the average cosine similarity between a node and its neighbor nodes. Then we sample random sets of the same size for each node containing the same number of random neighbors. We repeat 1000 times for each node to obtain a measure of how frequently random neighbor sets are as similar as the true sets. Using a generous cutoff, we ask for what fraction of the nodes are fewer than 10% of the random sets at least as similar as the true neighbors, summarized in Table 2.

**Table 2:** evaluation of concordance of language model and graph structure.

|  |  |
| --- | --- |
| Language model | Fraction of nodes concording with OMOP graph |
| biobert | 0.5830421074 |
| biogpt | 0.9613970588 |
| biomedbert | 0.5833622652 |
| biomegatron | 0.6129551871 |
| bluebert | 0.6386493557 |
| clinicalkb | 0.672130339 |
| sapbert | ? |

### Enriching causal inference models with knowledge representations

We develop a model to integrate a person’s demographic information, their history of the important variables, and the knowledge representation of each of these variables. To this end, electronic health records are converted into a graph. Each of people, drugs, conditions, etc, become nodes in this graph, and the embeddings from the language models comprise the features of these nodes. Each person node is connected to the variables in their medical history. Person nodes also have features which derive from the All of Us observation table including both survey data, and health record summaries (i.e., “How often have you felt depressed in the past two weeks?”. After filtering, the number of people’s features is 114.

To accommodate both person node features and medical code features, we use a heterogenous graph neural network. We train a heterogeneous convolution operation corresponding to each edge type to create a person’s represention combining their original node features with the connected medical codes. Finally, that person representation is passed through an MLP predicting propensity.

## Results

### Overview of target trials emulated and confounding associations

We emulate two active comparator target trials for drug safety, using the All of Us cohort (details in Methods). These target trials were selected by identifying the most common medications used in this population, then identifying drugs with at least two distinct indications in different body systems. We make the assumption that associations due to known confounding are more likely than any unreported true causal effect of either drug on the outcome.

In Comparison 1 (C1) we compare metopropolol versus sumatripan for effect on myocardial infarction. Metoprolol is a beta blocker used for cardiac disorders, but it is also approved for migraine headaches. Our first target trial compares the effect of metoprolol against another migraine medication, sumatriptan, asking whether either is associated with risk of myocardial infarction. We expect that metoprolol will be much more common in people at risk of myocardial infarction, making adjustment for confounding health risk a difficult task.

In Comparison 2 (C2), we include a more realistic target trial comparing two antidepressants to each other. We assess drug effect on chronic obstructive pulmonary disease (COPD). As we have found in our previous work, antidepressants have diverse therapeutic uses, making such active comparator trials subject to confounding(Kleper et al. 2025; Lalagkas and Melamed 2025b; Melamed 2020). We compare bupropion, an antidepressant with a secondary indication for smoking cessation, against sertraline, an antidepressant without such a secondary indication. As, again, only one of the two drugs is associated with a history of smoking, we expect a baseline association with COPD. Our target trial emulation approach is described in Table 1.

### Analysis of effect of potential confounding bias in causal inference models

We perform the active comparator analysis described above first without adjustment for potential confounding health history. As a result, we obtain the crude (unadjusted) risk ratio of 6.1 for C1, and 1.2 for C2 (Figure 1A). We also obtain bootstrap confidence intervals for each risk ratio (see Methods). While the interval for C2 overlaps the null, the interval for C1 does not.

**Figure 1:**
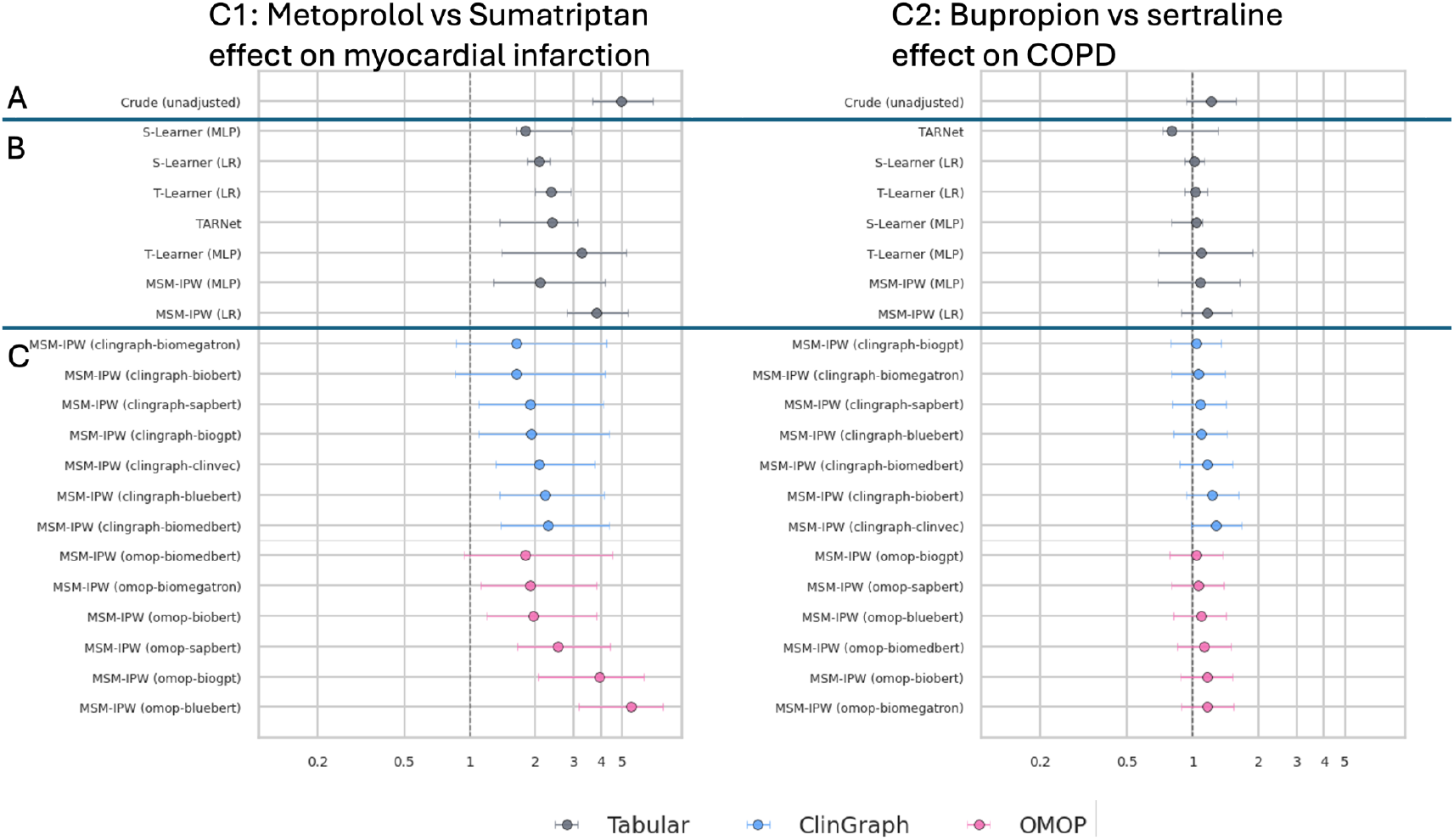
Emulation of two target trials using a variety of causal inference methods to obtain the average causal effect. **A.** Unadjusted effect. **B.** Potential confounders are encoded in a tabular format. **C.** Encoding confounders using language models (each rows) and knowledge graphs (ClinGraph in blue, OMOP concept relation table in red).

Next, we assess whether adjusting for covariates can ameliorate the biased association. We encode each confounder as a one-hot column in a tabular format, and we use these representations as input for the methods for causal inference. We implement a range of methods for counterfactual inference, which predicts the outcome as a function of exposure and treatment history. These methods obtain a counterfactual estimate of the outcome under either treatment, which can be compared to obtain an estimated average treatment effect. The methods vary both in terms of their learning algorithm (regression or deep learning) and in terms of the architecture of the learning task. T-learners learn a model for each of the two treatment cohorts, while S-learners learn one model incorporating all cohort data(Künzel et al. 2019). TARNet incorporates elements from both designs(Shalit et al. 2016). We also implement methods to learn the probability (propensity) for treatment, and use this to perform weighted regression identifying an estimated treatment effect(Austin 2011; Hernán and Robins 2024).

In Figure 1B we compare the effect estimates for C1 (left) and C2 (right). As compared to the baseline crude effect, the effect sizes appear to be most greatly reduced by S-learner and T-learners. While in C2 it appears that tabular data may suffice to remove the confounding association, in C1 the risk ratio generally remains above 2 in all cases. Next, we ask whether medical knowledge encodings can improve the results.

### Curation of foundation models and evaluation of consistency of foundation models with health concept knowledge graphs

As mentioned above, the tabular models simply represent each covariate as a one-hot. This reduces model complexity but does not leverage knowledge about the meaning of each variable. To encode this knowledge, we use both large language models learned from medical records and biomedical text, as well as knowledge graphs reprenting the relationship between these concepts. We curate a set of foundation models available from HuggingFace or direct download from the authors websites. These include: biobert, biogpt, biomedbert, biomegatron, bluebert, clinicalkb, and sapbert. We use each model to obtain an embedding representation *e*_*i*,_for each confounding concept *i* (see Methods).

We experiment both with the utility of these embeddings, and with combining the embeddings with knowledge graphs We ask whether the information in the knowledge graphs and embeddings is consistent as described in Methods, making a score for each node describing if the embedding of that node concords with the relations in the knowledge graph. We find that more than half of the nodes in all cases have concordant representations, with biogpt greatly exceeding the rest of the models.

### Enrichment with of causal inference models with knowledge representations

We experiment with multiple ways of incorporating these embeddings into the causal inference models. First, we take the average embedding for each individual, comprising the set all confounder codes *C* in their medical history (or 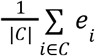). This replaces the one-hot representation for each person used in the tabular models, and then the model is trained as before. Secondly, we encode a person’s history of confounders into a knowledge graph, as described above, also encoding the patient’s baseline survey data into a feature vector for that person node. This allows us to represent each person as a convolution of their baseline information and the medical codes they connect to. Again, we use this model to find propensity for drug taking. The graph models have slightly higher AUC of predicting propensity than the baseline models (Figure 2). For both C1 and C2, the ATE is moved closer to the null (Figure 1C), though confidence intervals remain wide. This may be due to the larger number of parameters in these models.

**Figure 2:**
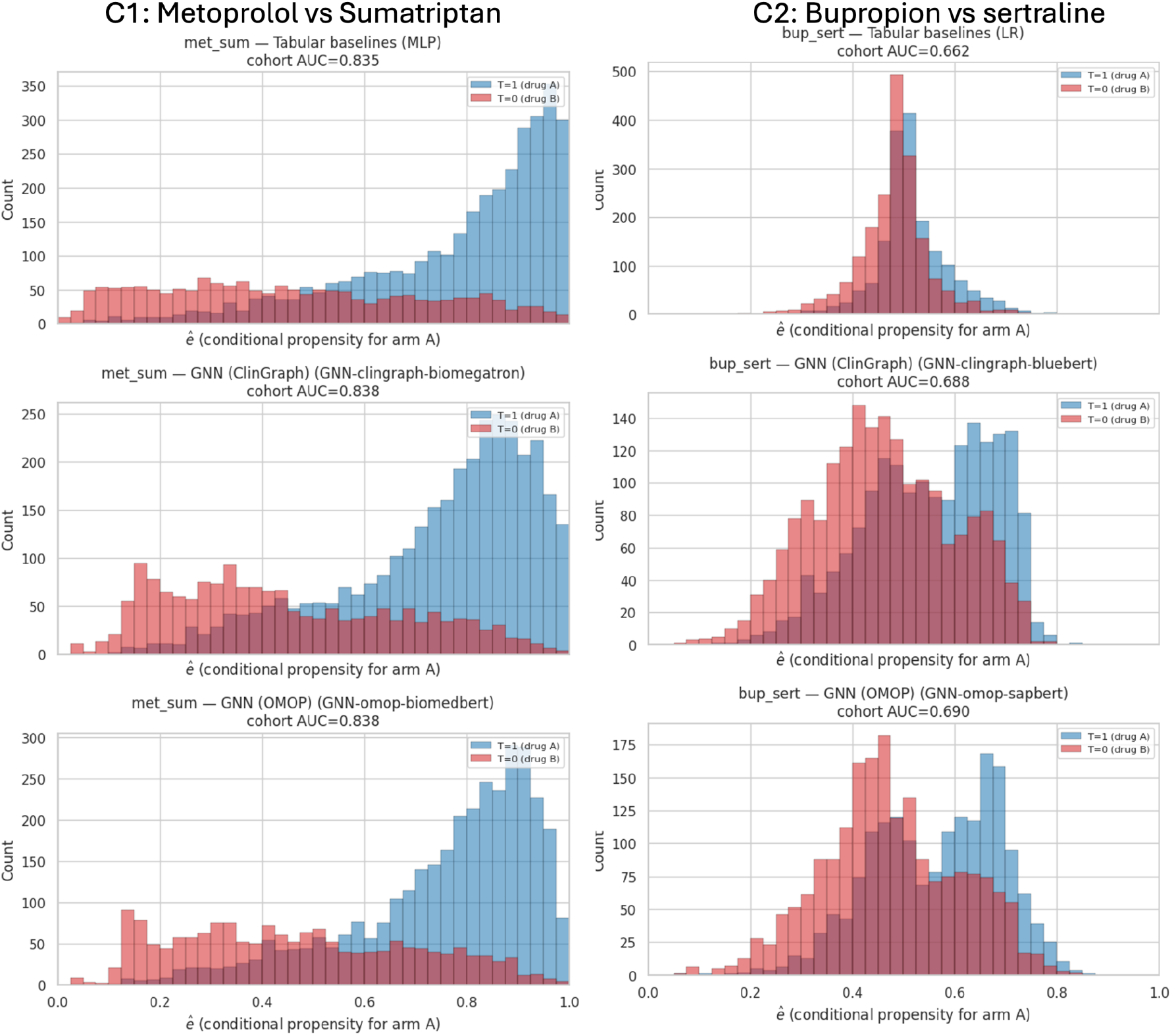
Comparison of propensity models estimates with tabular data (top) and language models and knowledge graphs (bottom two rows)

## Discussion

Many recent studies put forward new machine learning models for causal inference, new foundation models for biomedical data, and also health knowledgh graphs. But, rarely are the empirical utility of these models assessed for real-world tasks(Wornow et al. 2023). Here we specifically evaluate their potential to mitigate confounding in difficult case studies of confounding.

While these case studies may not be of particular interest, our goal is to assess the ability to overcome known confounding. These examples are also not of course the only examples of confounding. We expect that a high-throughput target trial emulation testing many drug effects would uncover more examples of confounding. Because our target trial emulation is able to be expanded to assess a range of phenotype outcomes, or a range of drug comparisons, such a systematic drug-wide analysis could help us better understand the weaknesses of various methods(Kleper et al. 2025; Ioannidis 2016).

Our analysis is the first, to our knowledge, to focus specifically on likely confounding rather than likely null effects. That makes it useful for identifying areas of remaining improvement needed for models. It can be used as a test case for others developing foundation models or methods for causal inference, particularly for health data.

## Data Availability

All data are available from the All of Us research portal.

https://researchallofus.org

